# Do Covid-19 patients needing ICU admission have worse 6 months follow up outcomes when compared with hospitalized non-ICU patients? A prospective cohort study

**DOI:** 10.1101/2021.08.17.21262177

**Authors:** Süleyman Yıldırım, Seher Susam, Pınar Cimen, Sena Yapicioglu, Onur Sunecli, Özlem Ediboglu, Cenk Kirakli

## Abstract

**Introduction:** Studies focus on pathogenesis, clinical manifestations, and complications during the early phase of the coronavirus disease-19 (COVID-19). Long-term outcomes of COVID-19 patients who discharge intensive care unit (ICU) are unclear.

**Objectives:** We investigated the effect of COVID-19 on lung structure, pulmonary functional, exercise capacity and quality of life in patients discharge from ICU and medical ward.

**Methods:** A prospective single-centre study conducted in PCR confirmed COVID-19 patients who has been discharged from University of Health Sciences, Dr. Suat Seren Chest Disease and Thoracic Surgery Teaching and Research Hospital between 15 January and 5 March 2021. Patients who followed up for more than 48 hours in ICU and more than 72 hours in medical ward were included the study. Computed tomography scores, pulmonary functional tests (PFT), 6-min walking distance and health related quality of life by SF-36 were compared between ICU and medical ward patients at 6 months after discharge.

**Results:** Seventy patients were included final analyses and 31 of them discharged from ICU. ICU patients had higher CT scores than non-ICU patients at admission (17 vs 11) and follow up visit (6 vs 0). Two-three of ICU patients had at least one abnormal finding at control CT. Advanced age (OR 1.08, 95% CI 1.02-1.15) and higher CT score at admission (OR 1.13, 95% CI 1.01-1.27) were risk factors for having radiological abnormalities at control CT.

**Conclusion:** A number of COVID-19 survivors especially with severe disease could not fully recover after 6 months of hospital discharge.

## Introduction

Coronavirus disease-19 (COVID-19), caused by severe acute respiratory syndrome coronavirus-2 (SARS-CoV-2), has affected over 150 million people around the world as of April 28, 2021^1^.

The onset symptoms of COVID-19 are fever, fatigue, shortness of breath, and a dry cough^2,3^. Typically chest tomography findings are peripheral, subpleural ground-glass opacities, bilaterally patchy shadows^2,4,5^ and chest computed tomography (CT) findings are related to disease severity^6,7^. Although most cases were classified as mild, 14% of cases were severe and 5% of them were critical, requiring intensive care unit (ICU) admission^8^. Studies focus on pathogenesis, clinical manifestations, and complications during the early phase of the disease^9–13^, but long-term outcomes still remain unclear.

Some of the patients recovered completely, but some patients were unable to reach their former health status, despite a long recovery period. Symptoms such as fatigue and dyspnea persist in half of the patients who were discharged from hospital^14,15^. CT findings are reversible in the majority of COVID-19 patients^16^. However, data on improvement of CT findings in ICU patients who have higher CT scores are lacking. Complete recovery may take a long time in mild and moderate cases as well as in severe patients who require ICU admission. More studies are needed on long-term outcomes in the post-COVID period^17^, especially in ICU patients.

The aim of this study was to evaluate the long-term effects of COVID-19 on lung structures, pulmonary functions, exercise capacity and quality of life in discharged ICU patients and compare these findings with hospitalized non-ICU patients.

## Methods

This is a single-centre, prospective cohort study performed between 15 January and 5 March 2021 at the University of Health Sciences, Dr. Suat Seren Chest Disease and Thoracic Surgery Teaching and Research Hospital. This is a tertiary hospital specializing in pulmonary diseases and has been designated for patients with COVID-19 since March 2020. Our study was approved by the ethics committee of University of Health Sciences, Dr. Suat Seren Chest Disease and Thoracic Surgery Teaching and Research Hospital (ethical approval number: 19-28.09.2020). Written informed consent was obtained from all participants. This study was registered on clinicaltrials.gov under the number NCT04715919.

## Patients

Patients who were followed up on for more than 48 hours in ICU and more than 72 hours in medical wards due to COVID-19 between March 19 and September 1, 2020 were included in the study. We excluded the following patients, (1) those who have neurodegenerative diseases, (2) those who were readmitted to the hospital due to any other conditions, (3) those with impaired movement due to physical disabilities. Patients were included in the study for at least 6 months, after discharge from the hospital.

The diagnosis of COVID-19 was based on World Health Organization (WHO) interim guidance^18^. Antiviral treatment against severe acute respiratory syndrome coronavirus 2 (SARS-CoV-2) was given according to Turkish Ministry of Health COVID-19 Guidance^19^.

## Procedures

Clinical data including demographic characteristics (age, gender, smoking status), antiviral treatment (hydroxychloroquine, favipiravir, convalescent plasma, steroids), chest tomography results and complication development during hospitalization were obtained from the hospital electronic record system. Patients were classified into 2 groups: ICU patients (followed in the ICU due to COVID-19) and non-ICU patients (hospitalized due to COVID-19 but followed in the pulmonary ward).

Participants were invited to the follow-up visit, by healthcare professionals, by telephone. All participants were consulted face to face by an investigator; asked to complete a questionnaire to assess their health status (Short Form-36, SF-36); and asked for persistent symptoms such as dyspnea, fatigue, muscle weakness etc. A trained physiotherapist performed a 6-min walking test to assess functional exercise capacity. The pulmonary function test was performed in the Pulmonary Functional Centre in University of Health Sciences, Dr. Suat Seren Chest Disease and Thoracic Surgery Teaching and Research Hospital according to American Thoracic Society and European Respiratory Society spirometry standardizations^20^.

Chest high-resolution computed tomography (HRCT) was performed at the end-inspiration in supine position, 1.25 mm section thickness and 0.625 mm reconstruction with high resolution. An experienced radiologist cross-compared HRCT images during hospital stay and follow-up HRCT images. If a participant had more than one HRCT, final chest images were included in the comparison. Both lungs were divided into 5 lobes in accordance with normal anatomical structure. Each lung lobe was given a score according to the following criteria; 0, no involvement; 1, less than 5% involvement; 2, 5-25% involvement; 3, 25-50% involvement; 4, 50-75% involvement; 5, more than 75% involvement. The total CT score was calculated semi-quantitatively with the sum of the scores of the five lobes^21^.

The SF-36 test is a 36-item self-reported survey of quality of life. SF-36 contains eight categories that assess physical functioning, social functioning, role limitation due to physical and emotional problems, general and mental health, bodily pain and vitality. Each category is scored from 0 (worst) to 100 (best) with higher scores showing better quality of life^22^. The translated and validated version of SF-36 was used for the study^23^.

The primary outcome was the percentage of patients with lung involvement in the 6-month follow up CT scan.

Secondary outcomes were exercise capacity (distance of 6-min walking test), pulmonary function tests and health state scores at the follow up visit.

## Statistical Analyses

Data are expressed as median (IQR) or number (%) where appropriate. Continuous data are compared with the Mann Whitney U test, and categorical data with the Chi square test. According to a study performed in severe acute respiratory syndrome (SARS), 30% of patients showed abnormal radiological findings at the 6 month follow up^24^. We also hypothesized that Covid 19 patients requiring ICU admission would have a 2-fold increase in the lung involvement at the 6 months follow up CT scan when compared with non-ICU patients. Assuming that 30% of non-ICU patients and 60% of ICU patients will have lung involvement in the 6-month follow-up CT, with a 5% type 1 error and 80% power, 40 patients in each group were needed for the analysis. A p value of <0.05 was considered as significant.

## Results

A total of 269 COVID-19 patients were discharged from our hospital between 1 April and 1 September 2020 and 70 patients were included in the study (Figure 1). The demographic and clinical characteristics of the participants are shown in table 1. Median age of participants was 56 years, and 60 (75%) of them were male. Most common comorbid diseases were hypertension (40%) and DM (33%). Thirty-one patients (44%) were admitted to the ICU and the median length of ICU stay was 9 days. Median length of hospital stay was 12 days and the time from the onset of symptoms to follow-up visit was 198 days.

**Table 1.** Demographic and clinical characteristics of the patients

| Patient characteristics | ICU patients (n=31) | Non-ICU patients (n=39) | p |
| --- | --- | --- | --- |
| Age, years | 59 (48-65) | 56 (48-61) | 0.40 |
| Male gender, n (%) | 26 (84) | 24 (62) | 0.04 |
| BMI, kg/m<sup>2</sup> | 31 (29-35) | 30 (25-34) | 0.20 |
| Smoking Status, n (%) |  |  |  |
| None | 14 (45) | 27 (69) |  |
| Active Smoker | 0 (0) | 2 (5) |  |
| Former Smoker | 17 (55) | 10 (26) |  |
| Smoking time, years | 30 (10-40) | 15 (13-25) | 0.25 |
| Comorbidities, n (%) |  |  |  |
| COPD | 5 (16) | 2 (5) | 0.13 |
| Hypertension | 16 (52) | 12 (31) | 0.07 |
| DM | 13 (42) | 10 (26) | 0.15 |
| CAD | 3 (10) | 2 (5) | 0.46 |
| CHF | 1 (3) | 1 (2) | 0.86 |
| Malignancy | 0 (0) | 3 (8) | 0.11 |
| Charlson Comorbidity Index | 1 (1-2) | 1 (1-2) | 0.16 |
| Disease Severity |  |  |  |
| 1- No O<sub>2</sub> Therapy | 0 (0) | 18 (46) | <0.001 |
| 2- Only O<sub>2</sub> Therapy | 3 (10) | 21 (54) |  |
| 3- NIV or HFNC | 23 (74) | 0 (0) |  |
| 4- IMV | 5 (16) | 0 (0) |  |
| APACHE-2 Score | 11 (8-15) | - |  |
| Treatment, n (%) |  |  |  |
| Hydroxychloroquine | 21 (68) | 33 (85) | 0.09 |
| Favipravir | 31 (100) | 19 (49) | <0.001 |
| Corticosteroid | 26 (83) | 3 (8) | <0.001 |
| Convalescent plasma | 12 (39) | 0 (0) | <0.001 |
| Tocilizumab | 5 (16) | 0 (0) | 0.009 |
| Antibiotics | 31 (100) | 37 (95) | 0.2 |
| Complications, n (%) |  |  |  |
| Acute renal failure | 4 (13) | 2 (5) | 0.39 |
| Hepatotoxicity | 9 (29) | 4 (10) | 0.045 |
| Sepsis | 1 (3) | 0 (0) | 0.26 |
| Length of hospital stay, days | 19 (14-28) | 8 (6-11) | <0.001 |
| Length of ICU stay, days | 9 (6-15) | - |  |
| Time from symptoms to follow up, days | 209 (189-219) | 190 (186-209) | 0.016 |

APACE-2, Acute Physiology and Chronic Health Evaluation; BMI, Body Mass Index; CHF, Congestive Heart Failure; COPD, Chronic Obstructive Pulmonary Disease; CAD, Coronary Arterial Disease; DM, Diabetes Mellitus; HFNO, High Flow Nasal Oxygen; ICU, intensive care unit IMV, Invasive Mechanical ventilation; NIV, Non-invasive Mechanical Ventilation Data are shown as n (%) and median (25<sup>th</sup> – 75<sup>th</sup> percentiles)

**Figure 1.**
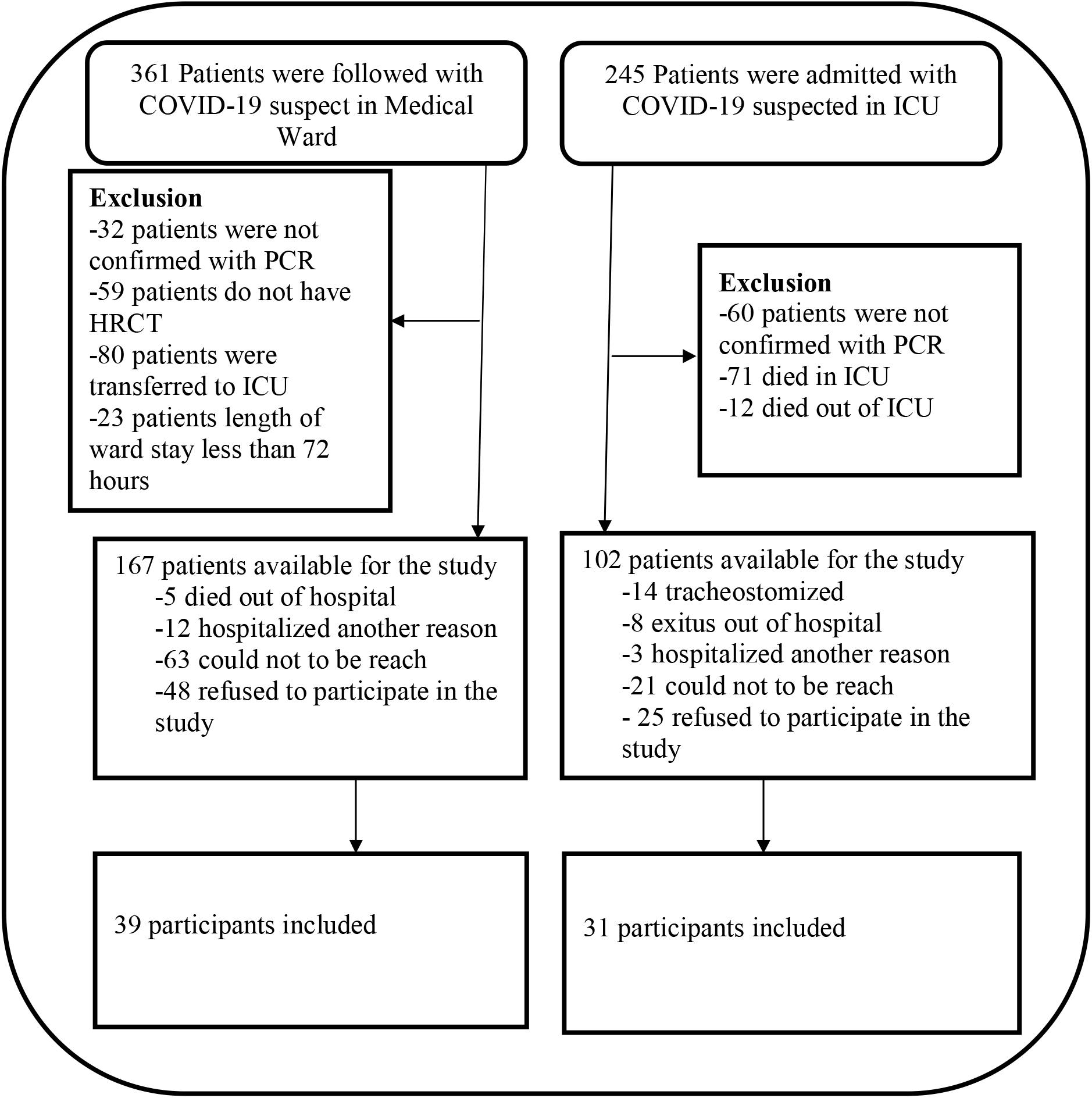
Flow chart of patients who admitted to hospital with suspect of COVID-19 during 19 March and 1 September 2020. COVID-19, Coronavirus Disease-19; HFNO, High Flow Nasal Oxygen; HRCT, High Resolution Computerized Tomography; ICU, Intensive Care Unit; IMV, Invasive Mechanical Ventilation; NIV, Non-invasive Mechanical Ventilation; PCR, Polymerase Chain Reaction

ICU patients had a higher median CT score at admission than non-ICU patients [17 (11-24) vs. 11 (10-15) respectively, p=0.002]. CT scores at the follow-up visit remained higher in ICU patients. All of the patients had at least one CT finding at admission, 20 ICU patients (65%) and 12 non-ICU patients (31%) had at least one CT finding in the follow-up CT (p=0.005) (Table 2). The most common CT finding was ground glass opacity (GGO) in the follow-up CT, followed by subpleural lines and irregular lines.

**Table 2.** Chest CT scores and findings at admission and 6-months follow up

| **CT findings** | **ICU patients**<br>**(n=31)** | **Non-ICU patients**<br>**(n=39)** | **p** |
| --- | --- | --- | --- |
| CT score at admission, points | 17 (11-24) | 11 (10-15) | 0.002 |
| Number of patients having at least one CT finding at admission, n % | 31 (100%) | 39 (100%) | 0.36 |
| CT score at 6 months follow-up visit | 6 (0-10) | 0 (0-3) | 0.001 |
| Number of patients having at least one CT finding at 6 months follow up visit, n % | 20 (65%) | 12 (31%) | 0.005 |

CT, Computerized Tomography; ICU, Intensive Care Unit Data are shown as n (%) and median (25<sup>th</sup> – 75<sup>th</sup> percentiles)

79% of participants had at least one persistent symptom. Individuals who were discharged from the ICU had a higher percentage of persistent symptoms, 90% vs 67% (p=0.033). Effort dyspnea was the most common persistent symptom followed by fatigue and muscle weakness (e–Table 1). Women had a higher percentage of persistent symptoms than men (e–Table 2).

A total of 65 participants (93%) completed the pulmonary function test (PFT) and 5 participants were unable to complete the test. Results of PFT are summarized at table 3. Forced vital capacity (FVC), peak expiratory flow (PEF) and peak inspiratory flow (PIF) were the most commonly affected parameters in ICU patients. PEF and PIF were also the two most commonly affected parameters in non-ICU patients. Higher CT scores at follow-up visits were found to be associated with impairment of PFT (Table 4).

**Table 3.** Results of pulmonary function tests, 6-MWD an quality of life scores at follow-up visit

| **Parameters** | **ICU patients**<br>**(n=31)** | **Non-ICU patients**<br>**(n=39)** | **p** |
| --- | --- | --- | --- |
| FVC, % of predicted | 85 (77-97) | 95 (85-104) | 0.73 |
| FEV1, % of predicted | 95 (84-103) | 99 (86-106) | 0.18 |
| FEV1/FVC, % of predicted | 112 (108-116) | 109 (102-114) | 0.94 |
| PIF, % of predicted | 96 (76-127) | 100 (64-132) | 0.98 |
| PEF, % of predicted | 90 (64-110) | 79 (68-96) | 0.42 |
| 6MWD, meters | 445 (346-515) | 461 (390-527) | 0.31 |
| FVC < 80% of predicted, n% | 9 (31%) | 5 (14%) | 0.09 |
| FEV1 < 80% of predicted, n% | 6 (21%) | 7 (19%) | 0.9 |
| FEV1/FVC < 80% of predicted, n% | 2 (7%) | 0 (0%) | 0.10 |
| PIF < 80% of predicted, n% | 8 (28%) | 12 (33%) | 0.61 |
| PEF < 80% of predicted, n% | 12 (41%) | 19 (49%) | 0.36 |
| 6MWD < 80% of mean distance | 13 (42%) | 11 (28%) | 0.22 |
| **SF-36 Categories** |  |  |  |
| Physical functioning | 80 (45-90) | 80 (65-95) | 0.48 |
| Social functioning | 50 (25-75) | 63 (38-75) | 0.23 |
| Role limitation due to physical problems | 50 (0-100) | 50 (25-100) | 0.16 |
| Role limitation due to emotional problems | 33 (0-100) | 67 (0-100) | 0.49 |
| General health | 65 (40-80) | 60 (45-80) | 0.79 |
| Mental health | 68 (48-84) | 68 (52-76) | 0.43 |
| Bodily pain | 88 (55-100) | 90 (58-100) | 0.81 |
| Vitality | 65 (40-80) | 60 (45-80) | 0.68 |

6MWD, 6-Minute Walking Distance; FEV1, Forced Expiratory Volume in One Second; FVC, Forced Vital Capacity; ICU, Intensive Care Unit; PEF, Peak Expiratory Flow; PIF, Peak Inspiratory Flow; SF-36, Short Form-36 Data are shown as n (%) and median (25<sup>th</sup> – 75<sup>th</sup> percentiles)

**Table 4.** Association between CT scores and pulmonary functional test impairment

|  | **FVC< 80% of predicted** | **FVC≥ 80% of predicted** | **p** |
| --- | --- | --- | --- |
| CT scores at admission | 11 (9-20) | 14 (10-17) | 0.78 |
| CT scores at 6 months follow-up | 7 (4-10) | 0 (0-4) | 0.007 |
|  | **FEV1< 80% of predicted** | **FEV1≥ 80% of predicted** |  |
| CT scores at admission | 10 (10-11) | 15 (11-18) | 0.11 |
| CT scores at 6 months follow-up | 5 (0-10) | 0 (0-5) | 0.036 |
|  | **FEV1/FVC< 80% of predicted** | **FEV1/FVC≥ 80% of predicted** |  |
| CT scores at admission | 6 (3-8) | 14 (10-18) | 0.024 |
| CT scores at 6 months follow-up | 5 (0-10) | 1 (0-8) | 0.80 |
|  | **PIF< 80% of predicted** | **PIF≥ 80% of predicted** |  |
| CT scores at admission | 11 (10-14) | 15 (11-18) | 0.07 |
| CT scores at 6 months follow-up | 3 (0-7) | 0 (0-9) | 0.55 |
|  | **PEF< 80% of predicted** | **PEF≥ 80% of predicted** |  |
| CT scores at admission | 11 (8-15) | 15 (12-19) | 0.006 |
| CT scores at 6 months follow-up | 2 (0-6) | 0 (0-8) | 0.19 |

CT, Computerized Tomography; FEV1, Forced Expiratory Volume in One Second; FVC, Forced Vital Capacity; ICU, Intensive Care Unit; PEF, Peak Expiratory Flow; PFT, Pulmonary Functional Test; PIF, Peak Inspiratory Flow
Data are shown as median (25<sup>th</sup> – 75<sup>th</sup> percentiles)

The median distance of 6-minute walk tests was similar in both groups; 445m in ICU patients, and 461 meters in non-ICU patients. When participants were stratified into age and weight, 13 participants (42%) in the ICU group, and 11 participants (28%) in the non-ICU group, were below 80% of the expected distance.

Assessment of quality of life, by SF-36, were similar in the two groups. Social functioning, role limitation due to physical and emotional problems were the most affected SF-36 categories (Table 3). The quality of life scores were lower in female participants than male participants (e–Table 3).

When the presence of ICU admission, age, gender, and CT score at admission were introduced into a logistic regression model, only patients age (OR 1.08, 95% CI 1.02-1.15) and higher CT score at admission (OR 1.13, 95% CI 1.01-1.27) were independent risk factors for having at least one radiological abnormality at a follow-up CT.

## Discussions

The main finding of this study was that the CT findings and symptoms (especially effort dyspnea, fatigue and muscle weakness) may not be totally resolved 6 months after the onset of symptoms in patients who require ICU admission. Also, some of these patients may encounter impaired pulmonary function tests and decreased exercise capacity. Impairment of quality of life was comparable between ICU and non-ICU patients.

Chest CT has been frequently used as a diagnostic tool during the COVID-19 outbreak and CT severity scores were related to disease severity^25^. In our study patients who were admitted to ICU had higher CT scores than patients admitted to the medical ward and this result was consistent with previous studies^25,26^. At the follow-up visit patients who were discharged from ICU had higher CT scores than patients who were discharged from the medical ward (6 vs 0). CT findings were totally resolved in most non-ICU patients, but most ICU patients had abnormal CT findings at the follow-up visit. In a previous study most of the CT findings were resolved in non-severe COVID-19 patients within 4 weeks after discharge^16,27^. However, as the severity of disease increases the recovery time may be longer. In a large cohort, patients with increased disease severity had higher CT scores at follow-up visits^28^. We found two-thirds of ICU patients had at least one of the CT findings at follow-up visits; irregular lines, subpleural lines and GGO were the most common patterns in chest CT. We also found advanced age and higher CT scores at admission were risk factors for having abnormal CT findings at the follow-up visit. Disease severity was found to be an independent risk factor for percentage change of CT score in the previous study^28^. Positive pressure ventilation or higher levels of fraction of inspired oxygen (FiO_2)_, which are frequently used in serious patients, may themselves cause lung damage and may inhibit complete recovery^29–31^.

We found PFT impairment was more frequent in individuals who were discharged from ICU. Patients with severe disease were more prone to PFT impairment at the early convalescence phase and long-term^27,28^. Patients with PFT impairment had higher CT scores at the follow-up visit despite these patients having had lower CT scores at admission (Table-4). It is difficult to associate changes in pulmonary function tests with COVID-19, since the baseline pulmonary function status of patients is not fully known. Nevertheless, patients with impaired PFT had higher scores at the follow-up CT, suggesting a relationship between partial improvement in CT findings and PFT impairment.

Median distance of 6-min walk was similar in both the patients discharged from ICU and from the medical ward, and these results are consistent with a previous study^28^. However, in half of the patients discharged from the ICU, when the patients were stratified into age and weight, the 6-minute walk test was found to be less than 80% of the expected value. Immobilization, severity of illness, and use of corticosteroids are risk factors for reduced exercise capacity^32^. ICU patients had these risk factors, thus reduced exercise capacity could be expected in these patients. Prolonged immobilization after hospital discharge and restrictions to prevent transmission, such as general curfew, may have limited the mobilization of these patients in the recovery phase.

We found 79% of participants had at least one persistent symptom and patients with severe disease had a higher percentage of persistent symptoms. The percentage of residual symptoms in COVID-19 varies from 49% to 79% in previous studies^14,28^. Effort dyspnea, fatigue and muscle weakness were the most common persistent symptoms in our and a previous study^28^.

Impairment of quality of life was observed in SF-36 categories, especially social functioning, role limitation due to physical and emotional problems. Impairment of quality of life was similar in both ICU and non-ICU patients. This result is consistent with long term follow up in SARS patients^24^. The percentage of residual symptoms and impairment of health status were significantly higher in female participants. Female survivors were more prone to depression and anxiety after the previous SARS outbreak^33^. Severity of disease and female gender were found to be risk factors for persistent psychological symptoms^28^. Not just disease-related causes, but also social restrictions (such as quarantine, and curfew to prevent spread of the disease), increased stress, anxiety, and depression in females^34^. Psychological distress, anxiety and depression may aggravate persistent symptoms and influence the impairment of quality of life.

This study has several limitations. This is a single-centre study so these results cannot be generalized to other centres. Although the desired number of patients could not be reached in the ICU patients group, the study has enough power (0.83) to test the difference between the two groups for the primary outcome. We could not measure diffusion of carbon monoxide (DLCO), which is frequently impaired in patients with SARS or COVID-19, due to technical reasons in our pulmonary functional centre. The baseline data of PFT and 6-min walking distance were unknown so we cannot directly associate COVID-19 and PFT or 6-min walking impairment. The fact that the majority of the participants were men may have affected the results especially the assessment of quality of life.

## Conclusion

This study is one of the first studies comparing the long-term outcomes of patients with COVID-19 who were admitted to ICU and medical wards. A number of COVID-19 survivors could not fully recover within 6 months of hospital discharge. Unresolved CT findings, impaired PFT, and decreased exercise capacity might be persistent in ICU patients even after 6 months. COVID-19 survivors especially with severe disease may have persistent lung injury, so they should be followed up for a long time.

## Data Availability

N/A

## Authors’ declaration

The authors declare that the submitted work is their own and that copyright has not been breached in seeking its publication. They confirm that the article is an original work, has not been published before, and is not being considered for publication elsewhere in its final form, in either printed or electronic media.

## Authors’ Contributions

Süleyman Yıldırım: literature search, data collection, study design, analysis of data, manuscript preparation, review of manuscript

Seher Susam: literature search, analysis of data, manuscript preparation, review of manuscript

Pınar Cimen: literature search, data collection, review of manuscript

Sena Yapicioglu: literature search, data collection, review of manuscript

Onur Sunecli: literature search, data collection, analysis of data, manuscript preparation, review of manuscript

Özlem Ediboglu: literature search, data collection, review of manuscript

Cenk Kirakli: literature search, data collection, study design, analysis of data, manuscript preparation, review of manuscript

