## Supplementary material for "Do Covid-19 patients needing ICU admission have worse 6 months follow up outcomes when compared with hospitalized non-ICU patients? A prospective cohort study": e-Table 1

e–Table 1. Number of patients having persistent symptoms in 6-months follow up

| **Symptoms** | **ICU**  **patients**  **(n=31)** | **Non-ICU**  **patients**  **(n=39)** | **p** |
| --- | --- | --- | --- |
| At least one persistent symptom | 28 (90%) | 27 (69%) | 0.033 |
| Fatigue | 12 (39%) | 14 (36%) | 0.80 |
| Muscle weakness | 8 (26%) | 8 (21%) | 0.60 |
| Cough | 1 (3%) | 3 (8%) | 0.42 |
| Dyspnea at rest | 7 (23%) | 7 (18%) | 0.63 |
| Exercise dyspnea | 21 (68%) | 20 (51%) | 0.16 |
| Taste/smell disorder | 1 (3%) | 2 (5%) | 0.69 |
| Headache | 1 (3%) | 4 (10%) | 0.25 |
| Sleep disorder | 6 (19%) | 5 (13%) | 0.45 |
| Hair loss | 5 (16%) | 4 (10%) | 0.46 |
| Diarrhea | 1 (3%) | 0 (0%) | 0.25 |
| Chest pain | 2 (6%) | 4 (10%) | 0.57 |
| Joint pain | 6 (19%) | 5 (13%) | 0.45 |
| Appetite loss | 0 (0%) | 1 (3%) | 0.36 |
| Vertigo | 3 (10%) | 3 (8%) | 0.76 |

ICU, Intensive care unit

Data are shown as n (%)
