## Supplementary material for "Do Covid-19 patients needing ICU admission have worse 6 months follow up outcomes when compared with hospitalized non-ICU patients? A prospective cohort study": e-Table 2

e–Table 2. Frequency of persistent symptoms at 6 months follow up according to gender

| **Symptoms** | **Male (n=50)** | **Female (n=20)** | **p** |
| --- | --- | --- | --- |
| At least one of following persistent symptom | 37 (74) | 18 (90) | 0.14 |
| Fatigue | 17 (34) | 9 (45) | 0.39 |
| Muscle weakness | 8 (16) | 8 (40) | 0.031 |
| Cough | 2 (4) | 2 (10) | 0.32 |
| Dyspnea at rest | 8 (16) | 6 (20) | 0.18 |
| Exercise dyspnea | 26 (52) | 15 (75) | 0.78 |
| Taste/smell disorder | 3 (6) | 0 (0) | 0.26 |
| Headache | 3 (6) | 2 (10) | 0.55 |
| Sleep disorder | 5 (10) | 6 (30) | 0.038 |
| Hair loss | 6 (12) | 3 (15) | 0.73 |
| Diarrhea | 1 (2) | 0 (0) | 0.52 |
| Chest pain | 6 (12) | 0 (0) | 0.10 |
| Joint pain | 5 (10) | 6 (30) | 0.38 |
| Appetite loss | 1 (2) | 0 (0) | 0.52 |
| Vertigo | 3 (6) | 3(15) | 0.22 |

Data are shown as n (%)
