## Supplementary material for "Do Covid-19 patients needing ICU admission have worse 6 months follow up outcomes when compared with hospitalized non-ICU patients? A prospective cohort study": e-Table 3

e–Table 3. Assessment of quality of life by gender

| **SF-36 Categories** | **Male**  **(n=50)** | **Female**  **(n=20)** | **p** |
| --- | --- | --- | --- |
| Physical functioning | 85 (75-90) | 58 (45-75) | <0.001 |
| Social functioning | 63 (37-87) | 50 (31-69) | 0.17 |
| Role limitation due to physical problems | 75 (25-100) | 0 (0-50) | 0.002 |
| Role limitation due to emotional problems | 67 (30-100) | 17 (0-67) | 0.011 |
| General health | 70 (50-85) | 45 (38-63) | 0.004 |
| Mental health | 70 (56-76) | 58 (40-68) | 0.026 |
| Bodily pain | 90 (78-100) | 59 (45-83) | <0.001 |
| Vitality | 45 (25-58) | 70 (50-85) | 0.002 |

ICU, Intensive Care Unit; SF-36, Short Form-36

Data are shown as median (25^th^ – 75^th^ percentiles)
